# Quality versus quantity of training datasets for artificial intelligence–based whole liver segmentation

**DOI:** 10.64898/2026.02.17.26346486

**Authors:** Austin Castelo, Caleb O’Connor, Aashish C. Gupta, Brian M. Anderson, McKell Woodland, Mais Altaie, Eugene J. Koay, Bruno C. Odisio, Tien T. Tang, Kristy K. Brock

## Abstract

Artificial intelligence (AI) based segmentation has many medical applications but limited curated datasets challenge model training; this study compares the impact of dataset annotation quality and quantity on whole liver AI segmentation performance. We obtained 3,089 abdominal computed tomography scans with whole-liver contours from MD Anderson Cancer Center (MDA) and a MICCAI challenge. A total of 249 scans were withheld for testing of which 30, MICCAI challenge data, were reserved for external validation. The remaining scans were divided into mixed-curation and highly-curated groups, randomly sampled into sub-datasets of various sizes, and used to train 3D nnU-Net segmentation models. Dice similarity coefficients (DSC), surface DSC with 2mm margins (SD 2mm), the 95th percentile of Hausdorff distance (HD95), and 2D axial slice DSC (Slice DSC) were used to evaluate model performance.

The highly curated, 244-scan model (DSC=0.971, SD 2mm=0.958, HD95=2.98mm) performed insignificantly different on 3D evaluation metrics to the mixed-curation 2,840-scan model (DSC=0.971 [*p*>.999], SD 2mm=0.958 [*p*>.999], HD95=2.87mm [p>.999]). The 710-scan mixed-curation (Slice DSC=0.929) significantly outperformed the highly curated, 244-scan model (Slice DSC=0.923 [*p*=0.012]) on the 30 external scans.

Highly curated datasets yielded equivalent performance to datasets that were a full order of magnitude larger. The benefits of larger, mixed-curation datasets are evidenced in model generalizability metrics and local improvements. In conclusion, tradeoffs between dataset quality and quantity for model training are nuanced and goal dependent.

## 1. Introduction

AI-based segmentation of medical images is a pivotal first step in many applications, including radiation treatment planning, large-scale longitudinal volume tracking, and intraoperative guidance. This has led to the rapid increase in the use of medical-image segmentation algorithms, with a 13.6% annual growth in related publications from 2010 to 2020 (Zhang et al., 2021). The rapid expansion of segmentation algorithms and the abundance of natural imaging datasets have created a desire to apply natural imaging segmentation approaches to medical imaging.

Natural image segmentation is built using large quantities of data, for example public benchmark datasets such as COCO (328,000 images) (Lin et al., 2014), Cityscapes (25,000 images) (Cordts et al., 2015), and ADE20K (20,000 images) (Zhou et al., 2017). Generating and curating these large, natural imaging datasets is relatively easy. In general, the underlying images are 2-dimensional (2D) representations of the physical world which allows anyone to perform image segmentation and annotation to generate ground truth without any additional training and at a low cost.

Medical image segmentation, however, has significant dataset creation and curation constraints. Data privacy concerns surrounding personal health information and data sharing agreements heavily limit the ability to gather and curate multi-institutional datasets. Ground-truth segmentation generation requires highly specialized medical knowledge, limiting dataset availability and increasing the cost of ground truth generation. More time is also needed to generate ground truth segmentations because of the complexity and size of 3D images. These constraints result in a steep tradeoff between quality and quantity, and how to best allocate resources is unclear. To the best of our knowledge, no study has fully explored the tradeoff between data quantity and quality in this context.

In this study, we compared the effects of dataset annotation quality and quantity on the performance of whole liver AI-based segmentation models using the nnU-Net architecture (Isensee et al., 2021). We trained segmentation models with mixed-curation and highly curated datasets, with sizes ranging from 56 to 2,840 scans, and evaluated the performance of these models. In summary the contributions of this paper are:

- evaluation of the effect of data quantity during training on medical imaging segmentation performance with the use case of liver segmentation of computed tomography (CT) images and
- evaluation of the effect of training data quality on medical imaging segmentation performance with the use case of liver segmentation of CT images.

## 2. Methods

### 2.1. Data

This section describes the data used within the highly curated and mixed-curation training datasets and the test dataset. Figure 1 portrays a flow chart of the relationship between the full 3,089 CT scans and these datasets, while Table 1 summarizes the dataset sources.

**Figure 1.**
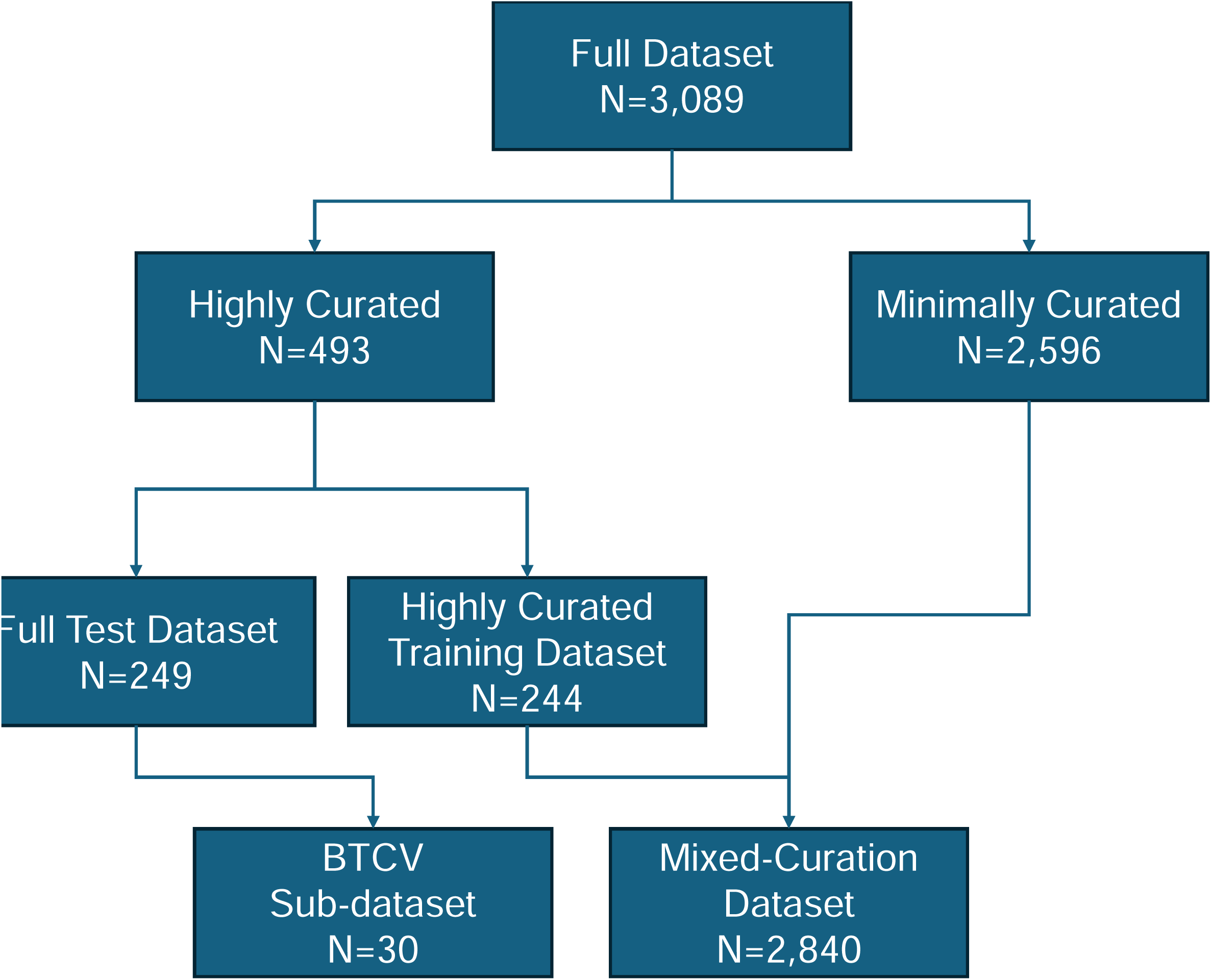
Flow chart of the relationship between the full dataset and the sub-datasets. Beyond the Cranial Vault (BTCV)

**Table 1.** Table of dataset sources used for the full dataset with the number of scans and description of the data.

| Source | Number of Scans | Description |
| --- | --- | --- |
| COVER-ALL | 257 scans | Liver ablation clinical trial data, contains planning and intra-operative CT imaging. (Odisio et al., 2025) |
| Anderson et al. | 205 scans | Internal dataset used by Anderson et al. (2021) contains contrast enhanced and non-contrast CT imaging. |
| BTCV | 30 scans | Multi-Atlas Labeling Beyond the Cranial Vault challenge data. (data : [https://www.synapse.org/ - !Synapse:syn3193805/wiki/89480](https://www.synapse.org/#!Synapse:syn3193805/wiki/89480)) |
| Internal Retrospective | 2,596 scans | Minimally curated retrospective data containing a large variety of CT scan protocols and quality. |

#### 2.1.1. Highly curated dataset

We identified 493 highly curated CT scans defined as CTs with manual segmentations generated or validated by a board-certified interventional radiologist. We sourced 258 CT scans from the COVER-ALL trial [NCT04083378] (Odisio et al., 2025), 205 CT scans from the training, validation, and test datasets used by Anderson et al. (2021), and 30 CT scans from the Multi-Atlas Labeling Beyond the Cranial Vault Medical Image Computing and Computer Assisted Intervention (BTCV) challenge (data: https://www.synapse.org/-!Synapse:syn3193805/wiki/89480). Together, these datasets led to a comprehensive set of images that varied in presence of contrast, phase of contrast (portal-venous phase versus arterial phase), and time that the image was acquired (intraoperatively versus postoperatively). Furthermore, a subset of scans contained non-anatomical structures, such as ablation needles, stents, and postresection clips.

From the 493 CT scans in the highly curated dataset, we created a highly curated training dataset containing 244 CT scans; the remaining highly curated scans were set aside for testing. These 244 scans consisted of only institutional data, 76 CT scans from the COVER-ALL trial [NCT04083378] (Odisio et al., 2025) and 168 CT scans from the training and validation datasets used by Anderson et al. (2021).

#### 2.1.2. Mixed-curation dataset

The mixed-curation training dataset was comprised of the highly curated training dataset described above and 2,596 minimally curated CT scans. These 2,596 minimally curated CT scans were sourced from an internal research database using the criteria that the CT scan had an associated liver contour obtained from unvalidated sources (e.g. early AI models, trainee’s segmentations, etc.). As the first step of our minimal curation, we trained a 3D nnU-Net model using the mixed-curation training dataset of 2,840 CT scans. Using the cross-validation results from the 5 folds, we evaluated the Dice similarity coefficient (DSC) between the model’s prediction and the unvalidated ground-truth contours on the 2,596 scans. Due to the lack of curation of the mixed-curation dataset we expected mislabeled or poor contours to be included; therefore, we performed a baseline quality control by assuming poor ground truth contours would have low performance in DSC evaluations. Subsequently, we had a radiologist qualitatively evaluate all scans with a DSC below 0.90 (N=65 scans) and correct the contour when necessary. Using this approach, we included the minimally curated CT scans resulting in a total dataset size of 2,840 for the mixed-curation training dataset (244 CT scans from the highly curated training dataset and 2,596 CT scans from the minimally curated dataset).

#### 2.1.3 Training sub-datasets

To evaluate the effect of training dataset size on segmentation models, we randomly sampled the mixed-curation training dataset at the following additional scan quantities: 56 (2% of the mixed-curation training dataset), 142 (5%), 284 (10%), 710 (25%), 1,420 (50%), 2,130 (75%), and 2,840 (100%). The mixed-curation training datasets contained the following proportions of highly curated scans: 2/56-scans (4%), 15/142-scans (11%), 21/284-scans (7%), 59/710-scans (8%), 119/1420-scans (8%), 182/2130-scans (9%), 244/2840-scans (9%). In addition, we randomly divided the highly curated training dataset into matched-size sub-datasets: 56 scans, 142 scans, and 244 scans (all the highly curated scans).

Mixed-curation training data may lead to a large variance in model results, especially when using small training datasets. To measure this variance, we resampled seven additional sub-datasets of the 56 scans from the mixed-curation training dataset for a total of eight 56-scan mixed-curation datasets. The models trained on these eight datasets were designated as Models 1-8, respectively.

#### 2.1.4 Test dataset

In total, we withheld 249 CT scans from the highly curated dataset for testing. There was no patient overlap with any training datasets and all highly curated external data was included. This was done to ensure that only the highest quality contours were evaluated and that they represented a diverse population of patients and acquisition techniques. Of the 249 scans withheld for testing, 30 scans were from the BTCV challenge, 182 scans were from the COVER-ALL trial (Odisio et al., 2025), and 37 scans were from Anderson et al. (2021) test dataset. Of the 219 internal scans (COVER-ALL and Anderson datasets), 54 intraoperative scans contained ablation probes and their associated artifacts. We evaluated the results on the full test set (249 CT scans) and the external BTCV challenge data to assess model generalizability by evaluating the model on external datasets not included in training.

### 2.2. Segmentation

An nnU-Net model was used in this study because of its state-of-the-art performance, automated configuration pipeline, and wide spread use to ensure our results have impact on the community (repository: https://github.com/MIC-DKFZ/nnUNet)(Isensee et al., 2021; Roy et al., 2023). The automatic configuration allowed us to simulate the hyperparameter tuning that would occur when training the model with our sub-datasets, removing any potential bias that only performing a hyperparameter search on the full dataset would introduce. Models 1-8 were trained using a default nnU-Net model utilizing the 3d_fullres configuration. The 9 remaining training sub-datasets were trained using nnU-Net models with an increased batches per epoch from the default 250 batches/epoch to 1000 batches/epoch. The highest performing model in Models 1-8 was retrained with 1000 batches/epoch for comparison with other models. All models used the 3d_fullres configuration using five Nvidia A100 graphic processing units (GPUs), with each model taking approximately 18 hours to train. For reference, Table 2 shows the mixed-curation and highly curated models with descriptions of the datasets on which each was trained.

**Table 2.** Table showing the minimally curated and highly curated model names and a description of the training dataset.

|  | Model | Description |
| --- | --- | --- |
| **Mixed-Curation models** | Models 1-8 | 2% (56 scans) random samples of mixed-curation training dataset |
|  | 56-scan | Model 1, best performing 2% random sample of mixed-curation training dataset |
|  | 142-scan | 5% random sample of mixed-curation training dataset |
|  | 284-scan | 10% random sample of mixed-curation training dataset |
|  | 710-scan | 25% random sample of mixed-curation training dataset |
|  | 1420-scan | 50% random sample of mixed-curation training dataset |
|  | 2130-scan | 75% random sample of mixed-curation training dataset |
|  | 2840-scan | 100% of mixed-curation training dataset |
| **Highly curated models** | 56-scan | Random sample of highly curated training dataset to match 56-scan minimally curated model |
|  | 142-scan | Random sample of highly curated training dataset to match 142-scan minimally curated model |
|  | 244-scan | Full highly curated training dataset |

### 2.3. Model evaluation

To comprehensively evaluate the models, we calculated the mean, standard deviation, median, and 1^st^ quartile of DSC (1); mean and standard deviation of surface DSC 2mm margin (SD 2mm) (2); mean, standard deviation, 1^st^ quartile, 5^th^ percentile, and mean within the 5^th^ percentile of 2D DSC on axial slices containing ground truth liver contours (Slice DSC); and mean and standard deviation of the 95th percentiles of the Hausdorff distance (HD95) (3) for each model. Owing to the liver’s large volume, segmentation performance on clinically relevant artifacts is often not fully captured by 3D volumetric DSC evaluations. Thus, we performed a 2D DSC evaluation of axial slices (Slice DSC), mean, 1^st^ quartile, 5^th^ percentile, and mean within the 5^th^ percentile, in addition to the raw number and proportion of slices with DSC of 0, <0.80, and <0.90 were reported to evaluate the worst performing slices for each model.

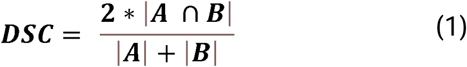

Equation 1. Where |A| and |B| are sets.

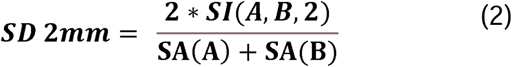

Equation 2. SI(A,B,T) is the Surface Intersection of surface A and surface B given tolerance T and SA(A) is the Surface Area of A.

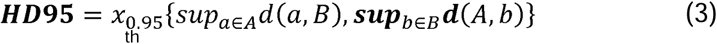

Equation 3. Where x_0.95_{X} is the 95 percentile of the set X and d(a, B) is the minimum distance between point a and surface B.

On the full test dataset and BTCV subset, we performed a Friedman test to determine if statistically significant performance differences were present across the DSC, SD 2mm, HD95, and Slice DSC results across all highly curated and mixed-curation models. If the Friedman test showed statistical significance, we performed a Dunn’s multiple comparisons test to compare model performance across the DSC, SD 2mm, HD95, and Slice DSC results of each model with those of the best case–scenario mixed-curation 56-scan model from Models 1-8 and the highly curated 244-scan model. We considered a *p* value less than 0.05 to represent a statistically significant difference between models for a given metric.

## 3. Results

The following sections showcase the evaluation results of the highly curated models and the mixed-curation models across the full test set and the external BTCV test set. We start with evaluating the variance of the small training datasets by evaluating Models 1-8 on the full test dataset. After which, the results of the remaining mixed-curation and highly curated models on the full test set will be explored following which the external BTCV sub-dataset will be explored.

### 3.1. Variance of small training datasets

Models 1-8 (trained on the random samples of 2% of the mixed-curation training dataset) had an average mean DSC of 0.960 (Figure 2). Model 1 showed the highest performance with a mean DSC = 0.964 and Model 8 showed the worst performance with a mean DSC = 0.955. The two poorest performing models, Models 4 and 8, showed a one-tailed distribution: the bottom 25th percentile had reduced performance owing to outlier scans, such as those containing metal artifacts, not being included in these randomly selected 56-scan sub-datasets. We used Model 1 as the mixed-curation 56-scan model in all further comparisons due to it showing peak performance across Models 1-8. (Figure 2)

**Figure 2.**
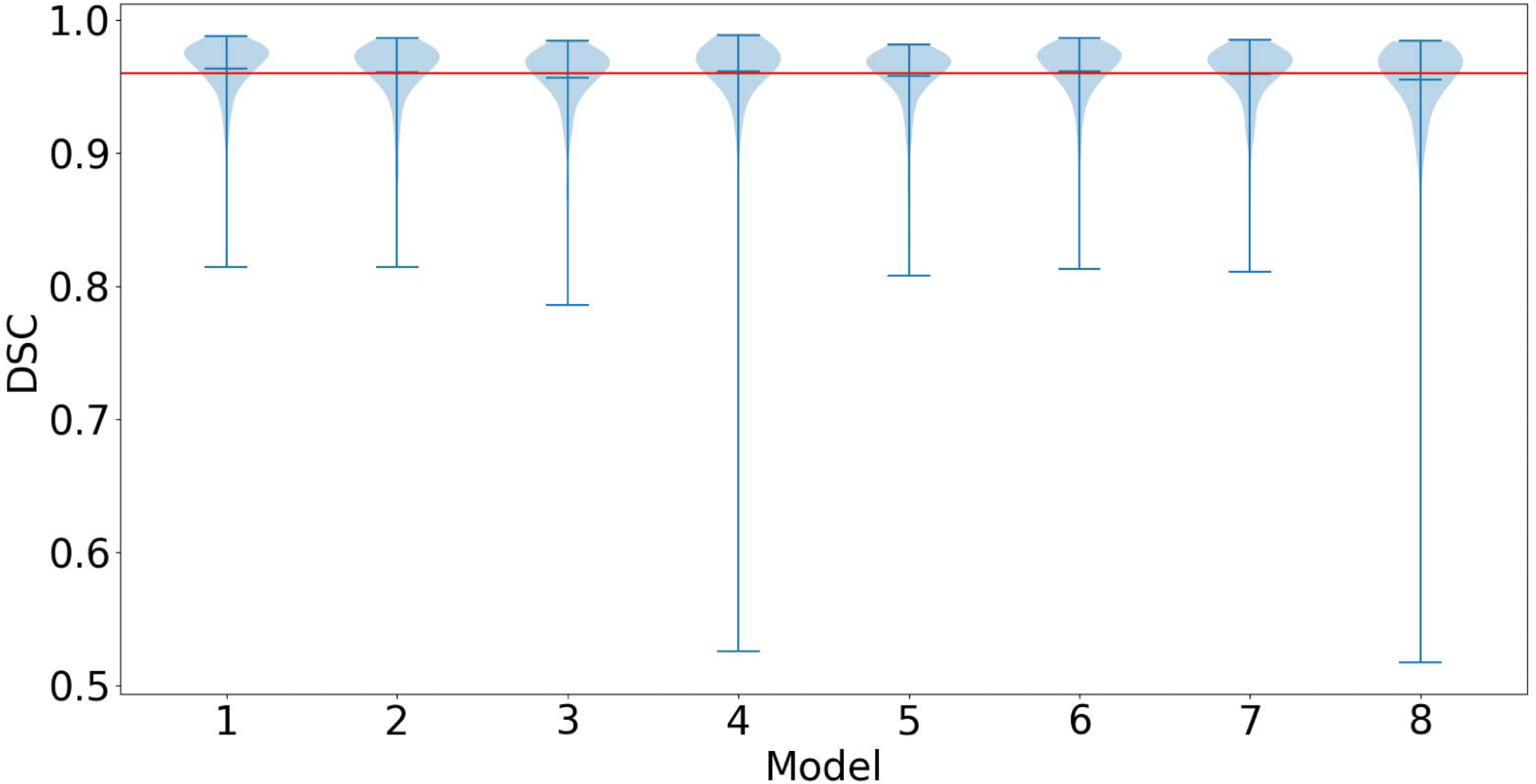
Violin plot comparing the Dice similarity coefficient (DSC) of 8 mixed-curation 56-scan models. The maximum, mean, and minimum bars are plotted for each model. The red line represents the average mean DSC across all models (0.96)

### 3.2. Full test dataset

The mean DSC, median DSC, 1st quartile DSC, mean SD 2mm, and mean HD95 performance results of testing with the full test dataset for all models are shown in Table 3 with *p* values shown in Supplementary Table 1. We did not observe a large range in the mean DSC (0.961-0.971), median DSC (0.968-0.976), or 1st quartile DSC (0.955-0.968) across all models. Significant performance increase is shown in the mixed-curation models as the training dataset size increased from 56 to 142 scans: the mean DSC increased from 0.961 to 0.967 (*p*<.001), the mean HD95 decreased from 4.56mm to 3.18mm (*p*<.001) and mean SD 2mm increased from 0.936 to 0.950 (*p*<.001), respectively. As the mixed-curation dataset size increased above 710 scans to the complete 2840 scans, a plateau in performance measured by mean DSC was observed; the mean DSC fluctuated between 0.970 and 0.971. Minor gains in performance measured by mean SD 2mm and mean HD95 was observed beyond 710 scans; the mean SD 2mm increased from 0.956 to 0.958 and mean HD95 decreased from 3.06mm to 2.87mm.

**Table 3.** Model performance on the Full Test Set (n=249). * *p* < 0.05 when compared against the mixed-curation 56-scan model (Dunn’s multiple comparisons test), ^ p<0.05 when compared against the highly curated 244-scan model (Dunn’s multiple comparisons test). *P-values* shown in Supplementary Table 1. Underlined values denote highest performing model in a metric. Dice Similarity Coefficient (DSC), Standard Deviation (STD), Hausdorff Distance 95^th^ percentile (HD95), Surface Dice similarity coefficient with 2mm margin (SD 2mm)

|  | Mixed-Curation models |  |  |  |  |  |  | Highly curated models |  |  |
| --- | --- | --- | --- | --- | --- | --- | --- | --- | --- | --- |
|  | 56-scan | 142-scan | 284-scan | 710-scan | 1420-scan | 2130-scan | 2840-scan | 56-scan | 142-scan | 244-scan |
| Mean DSC (STD) | 0.961<br>(0.025)^ | 0.967<br>(0.019)*^ | 0.969<br>(0.019)*^ | <u>0.971</u><br>(0.017)*^ | 0.970<br>(0.019)* | <u>0.971</u><br>(0.018)* | <u>0.971</u><br>(0.018)* | 0.969<br>(0.021)*^ | <u>0.971</u><br>(0.017)* | <u>0.971</u><br>(0.019)* |
| Median DSC | 0.968 | 0.972 | 0.974 | <u>0.976</u> | 0.975 | <u>0.976</u> | 0.975 | 0.975 | <u>0.976</u> | <u>0.976</u> |
| 1st Quartile DSC | 0.955 | 0.964 | 0.965 | 0.966 | 0.966 | 0.966 | 0.967 | 0.965 | <u>0.968</u> | <u>0.968</u> |
| HD95 Mean (STD) | 4.56<br>(8.49)^ | 3.18<br>(4.71)*^ | 4.42<br>(18.7)*^ | 3.06<br>(6.97)* | 2.97<br>(5.93)* | 2.95<br>(6.19)* | <u>2.87</u><br>(5.39)* | 3.59<br>(7.99)*^ | 3.14<br>(6.11)* | 2.98<br>(4.60)* |
| SD 2mm Mean (STD) | 0.936<br>(0.062)^ | 0.950<br>(0.052)*^ | 0.953<br>(0.051)*^ | 0.956<br>(0.048)* | 0.957<br>(0.049)* | 0.957<br>(0.048)* | <u>0.958</u><br>(0.048)* | 0.952<br>(0.054)*^ | <u>0.958</u><br>(0.047)* | <u>0.958</u><br>(0.048)* |

The highly curated models performed equivalently to much larger mixed-curation datasets, as shown in Table 3. The highly curated 244-scan model showed non-significant differences over all mixed-curation models with dataset sizes utilizing 1,420 scans or greater across mean DSC, mean HD95, or mean SD 2mm (*p*>.999). No significant improvement was observed between the highly curated 244-scan model and the highly curated 142-scan model in mean DSC (*p*>.999), mean HD95 (*p*>.999), and mean SD 2mm (*p*>.999).

The 2D evaluation metrics for the full test set are shown in Table 4 with *p* values in Supplemental Table 2, including Slice DSC mean, 5^th^ percentile, 1^st^ quartile, mean of results <5^th^ percentile, and counts of slices with DSC of 0, <0.80, and <0.90. We observed performance increases in mean Slice DSC as dataset size increased for the mixed-curation datasets up to the 710-scan model and highly curated datasets up to the 142-scan model. The mixed-curation 710-scan model showed the highest performance in the percentage of slices with DSC of 0 and <0.80 with 0.42% and 2.21% respectively. Minor degradation in performance was observed as dataset size increased beyond 710-scans. The 244-scan highly curated model (Slice DSC = 0.954) performed significantly worse than the mix-curation 710-scan model (Slice DSC = 0.957 [*p*<.001]) and 1420-scan model (Slice DSC= 0.956 [*p*<.001]).

**Table 4.** Model performance on 2D slices in the Full Test Set (n=14,082). * *p* < 0.05 when compared against the mixed-curation 56-scan model (Dunn’s multiple comparisons test), ^ p<0.05 when compared against the highly curated 244-scan model (Dunn’s multiple comparisons test). *P-values* shown in Supplementary Table 2. Underlined values denote highest performing model in a metric.

|  | Mixed-Curation models |  |  |  |  |  |  | Highly curated models |  |  |
| --- | --- | --- | --- | --- | --- | --- | --- | --- | --- | --- |
|  | 56-scan | 142-scan | 284-scan | 710-scan | 1420-scan | 2130-scan | 2840-scan | 56-scan | 142-scan | 244-scan |
| Mean Slice DSC (STD) | 0.936<br>(0.142)^ | 0.946<br>(0.113)*^ | 0.953<br>(0.096)*^ | <u>0.957</u><br>(<u>0.084</u>)*^ | 0.956<br>(0.090)*^ | 0.956<br>(0.090)* | <u>0.957</u><br>(<u>0.088</u>)* | 0.948<br>(0.112)*^ | 0.954<br>(0.100)*^ | 0.954<br>(0.100)* |
| 5<sup>th</sup> Percentile Slice DSC | 0.815 | 0.852 | 0.867 | <u>0.883</u> | 0.875 | 0.878 | 0.878 | 0.846 | 0.867 | 0.870 |
| 1st Quartile Slice DSC | 0.951 | 0.955 | 0.958 | 0.959 | 0.960 | 0.960 | <u>0.961</u> | 0.958 | 0.960 | 0.960 |
| <5<sup>th</sup> Percentile Mean Slice DSC | 0.429 | 0.573 | 0.649 | <u>0.704</u> | 0.674 | 0.677 | 0.685 | 0.574 | 0.637 | 0.636 |
| Slice DSC =0 (%) | 215<br>(1.53%) | 119<br>(0.85%) | 79<br>(0.56%) | <u>59</u><br>(<u>0.42%</u>) | 66<br>(0.47%) | 70<br>(0.50%) | 64<br>(0.45%) | 120<br>(0.85%) | 102<br>(0.72%) | 89<br>(0.63%) |
| Slice DSC <0.8 (%) | 643<br>(4.57%) | 474<br>(3.37%) | 396<br>(2.81%) | <u>311</u><br>(<u>2.21%</u>) | 363<br>(2.58%) | 348<br>(2.47%) | 343<br>(2.44%) | 513<br>(3.64%) | 402<br>(2.85%) | 410<br>(2.91%) |
| Slice DSC <0.9 (%) | 1368<br>(9.71%) | 1199<br>(8.51%) | 1021<br>(7.25%) | 928<br>(6.59%) | 936<br>(6.65%) | 938<br>(6.66%) | <u>922</u><br>(<u>6.55%</u>) | 1163<br>(8.26%) | 1013<br>(7.19%) | 1027<br>(7.29%) |

**Table 5.** Model performance on BCTV Test Set (n=30). * p<0.05 Dunn’s multiple comparisons test against mixed-curation 56-scan model, ^ p<0.05 Dunn’s multiple comparisons test against highly curated 244-scan model. *P-values* shown in Supplementary Table 4. Underlined values denote highest performing model in a metric. Dice Similarity Coefficient (DSC), Standard Deviation (STD), Hausdorff Distance 95^th^ percentile (HD95), Surface Dice similarity coefficient with 2mm margin (SD 2mm)

|  | Mixed-Curation models |  |  |  |  |  |  | Highly curated models |  |  |
| --- | --- | --- | --- | --- | --- | --- | --- | --- | --- | --- |
|  | 56-scan | 142-scan | 284-scan | 710-scan | 1420-scan | 2130-scan | 2840-scan | 56-scan | 142-scan | 244-scan |
| Mean DSC (STD) | 0.960<br>(0.009)^ | <u>0.963</u><br>(<u>0.009</u>) | 0.961<br>(0.012)* | <u>0.963</u><br>(<u>0.010</u>)* | <u>0.963</u><br>(<u>0.009</u>)* | <u>0.963</u><br>(<u>0.009</u>)* | <u>0.963</u><br>(<u>0.009</u>)* | 0.960<br>(0.012)^ | <u>0.963</u><br>(<u>0.011</u>)* | <u>0.963</u><br>(<u>0.011</u>)* |
| Median DSC | 0.962 | 0.965 | 0.965 | <u>0.966</u> | 0.965 | 0.965 | 0.965 | 0.962 | 0.965 | <u>0.966</u> |
| 1st Quartile DSC | 0.955 | <u>0.962</u> | 0.960 | 0.960 | 0.959 | 0.960 | 0.960 | 0.957 | 0.958 | 0.958 |
| HD95 Mean (STD) | 9.57<br>(18.40)^ | 7.03<br>(11.20) | 17.3<br>(52.00) | 7.80<br>(18.70)* | 7.23<br>(15.30) | 7.34<br>(16.20)* | <u>6.80</u><br>(<u>13.70</u>) | 9.89<br>(20.10)^ | 8.19<br>(15.40) | 6.83<br>(9.37)* |
| SD 2mm Mean (STD) | 0.931<br>(0.024)^ | 0.935<br>(0.023) | 0.934<br>(0.031) | 0.937<br>(0.024)* | 0.938<br>(0.023) | 0.938<br>(0.022)* | 0.938<br>(0.022)* | 0.931<br>(0.024)^ | 0.939<br>(0.022)* | <u>0.940</u><br>(<u>0.021</u>)* |

### 3.3 Generalizability of the model: BTCV test sub-dataset

Testing with the BTCV test sub-dataset resulted in a decrease in performance in comparison to testing with the full test dataset for all models across all metrics (Table 7, Supplementary Tables 3).

Performance improved as dataset quantity increased in both the mixed-curation and highly curated models from 56 scans to 142 scans. As the dataset size increased above 142 scans for both highly curated and mixed-curation models, mean DSC performance plateaued. The highly curated 56-scan model showed no significant performance increase (*p*>.999) in any metric compared to the mixed-curation 56-scan model. The 244-scan highly curated models showed no statistically significant improvement in any metric compared to any mixed-curation model except for the 56-scan mixed-curation model, where the 244-scan highly curated model showed a significant improvement (*p*<.05) across all metrics.

In contrast to what was observed on the full test set, for the Slice DSC analysis of the BTCV test sub-dataset the mixed-curation dataset models had a higher performance in mean Slice DSC than their equivalent dataset size highly curated models (Table 6, Supplementary Table 2). When evaluated based on the Slice DSC, the 244-scan highly curated model’s performance was significantly worse (*p<*.02) than all mixed-curation models with datasets equal to or larger than 284 scans except for the 2840-scan mixed-curation model (p>.999). Peak performance was observed in the 710-scan mixed-curation model for mean Slice DSC (0.929), 5^th^ percentile Slice DSC (0.821), <5^th^ percentile mean Slice DSC (0.382), and percentage of slices with Slice DSC equal to 0 (2.04%).

**Table 6.** Model performance on 2D slices in the BCTV Test Set (n= 1,517). * *p* < 0.05 when compared against the mixed-curation 56-scan model (Dunn’s multiple comparisons test), ^ p<0.05 when compared against the highly curated 244-scan model (Dunn’s multiple comparisons test). *P-values* shown in Supplementary Table 2. Underlined values denote highest performing model in a metric.

|  | Mixed-Curation models |  |  |  |  |  |  | Highly curated models |  |  |
| --- | --- | --- | --- | --- | --- | --- | --- | --- | --- | --- |
|  | 56-scan | 142-scan | 284-scan | 710-scan | 1420-scan | 2130-scan | 2840-scan | 56-scan | 142-scan | 244-scan |
| Mean Slice DSC (STD) | 0.914<br>(0.179)^ | 0.923<br>(0.160)* | 0.926<br>(0.157)*^ | <u>0.929</u><br>(0.150)*^ | 0.927<br>(0.156)*^ | 0.927<br>(0.158)*^ | 0.927<br>(0.158)* | 0.909<br>(0.189)*^ | 0.918<br>(0.179)* | 0.923<br>(0.168)* |
| 5<sup>th</sup> Percentile Slice DSC | 0.768 | 0.776 | 0.810 | <u>0.821</u> | 0.819 | 0.813 | 0.813 | 0.632 | 0.734 | 0.78 |
| 1st Quartile Slice DSC | 0.943 | 0.947 | 0.946 | 0.948 | 0.948 | <u>0.949</u> | <u>0.949</u> | 0.941 | 0.946 | 0.947 |
| <5<sup>th</sup> Percentile Mean Slice DSC | 0.196 | 0.309 | 0.347 | <u>0.382</u> | 0.347 | 0.338 | 0.335 | 0.139 | 0.206 | 0.269 |
| Slice DSC =0 (%) | 44<br>(2.9%) | 33<br>(2.18%) | 35<br>(2.31%) | <u>31</u><br>(2.04%) | 33<br>(2.18%) | 36<br>(2.37%) | 34<br>(2.24%) | 52<br>(3.43%) | 49<br>(3.23%) | 40<br>(2.64%) |
| Slice DSC <0.8 (%) | 89<br>(5.87%) | 83<br>(5.47%) | 70<br>(4.61%) | 69<br>(4.55%) | <u>68</u><br>(4.48%) | 71<br>(4.68%) | 71<br>(4.68%) | 110<br>(7.25%) | 95<br>(6.26%) | 81<br>(5.34%) |
| Slice DSC <0.9 (%) | 173<br>(11.4%) | 170<br>(11.21%) | 157<br>(10.35%) | 152<br>(10.02%) | 147<br>(9.69%) | 148<br>(9.76%) | <u>144</u><br>(9.49%) | 199<br>(13.12%) | 174<br>(11.47%) | 167<br>(11.01%) |

## 4. Discussion

To our knowledge, this is the first study to comprehensively compare training dataset size and quality in whole-liver image segmentation. We demonstrated that for the creation of whole-liver segmentation models there are benefits to both approaches of prioritizing quality and quantity in dataset creation. Tested on our full test set with a variety of scans from within our institution and scans from a public challenge we illustrated near equivalent performance across both approaches. Examining the results on the BTCV sub-dataset, we showcased an advantage in generalizability of the larger mixed-curation models.

Results from 3D evaluation on the full test dataset showcased low variance in performance across all models and the value of highly curated data in data scarcity. Improved performance in DSC, HD95, and SD 2mm between the lowest performing model, 56-scan mixed-curation (DSC = 0.961, HD95 = 4.56mm, SD 2mm = 0.936), and the highest performing model, 2840-scan mixed curation (DSC = 0.971, HD95 = 2.87mm, SD 2mm = 0.958 *p*<.0001), resulted in statistical significance but did not appear to be clinically significant. The performance increase of the highly curated 56-scan model (DSC = 0.969, HD95 = 3.59, SD 2mm = 0.952) over the mixed-curation 56-scan model (DSC = 0.961, HD95 = 4.56, SD 2mm = 0.936, *p*<.0001) demonstrated the value of high curation in data scarce situations.

Results from 2D slice evaluation on the full test dataset diverge from the 3D evaluation results on the comparison of mixed-curation and highly curated models. While the performance plateaus remain the same at 710-scans for the mixed-curation and 142-scans for the highly curated models on 3D DSC, for Slice DSC the equivalent performance of the highly curated models to the larger mixed-curation models no longer holds. As seen between the plateau points, the 710-scan mixed-curation model resulted in a mean Slice DSC of 0.957 and the highly curated 142-scan model resulted in a mean Slice DSC of 0.954. This divergence from the 3D evaluation results in addition to the increase in low performance slice, Slice DSC = 0 in Table 4, suggest that while the two approaches of mixed-curation and high-curation reached equivalent 3D performance there is an improvement in the larger mixed-curation models over the high-curation on a slice-by-slice basis, reflecting the models ability to accurately identify subtle features.

Results from testing with the BTCV sub-dataset demonstrated how training dataset quantity affects a model’s generalizability to entirely new datasets. As expected, there is a performance decrease across all models and all metrics on the BTCV dataset compared to the full test dataset. Low variance in performance is noted in mean DSC with values ranging from 0.960 to 0.963.

Combined with the low variance in ranges of mean HD95 and mean SD 2mm, excluding the 56-scan models, of 8.19mm to 6.80mm and 0.935 to 0.940, respectively, no clear signal for preference of either curation method based on 3D is observed. Analysis of the Slice DSC metrics revealed that mixed-curation models consistently outperformed their size-matched highly curated counterparts, with none of the highly curated models attaining comparable or better scores.

Beyond the direct comparison of dataset quality and quantity, the approach described in Section 2.1.2 provides a practical framework for iterative dataset refinement during model development. By using model disagreement or low-performance cases as a trigger for targeted expert review, training can proceed with minimally curated data while selectively investing expert time where it is most impactful. This strategy enables progressive improvement of both dataset quality and model performance without requiring exhaustive manual curation upfront, offering a scalable pathway for maintaining and expanding large clinical training datasets.

Recent work has shown promising results in out-of-distribution detection utilizing feature extractions (Woodland et al., 2024) and generative models (Woodland et al., 2025), which could be used to improve the curation of high-quality training datasets by selecting an optimal sub-dataset to generate ground truths for. In the context of the results from these studies, we suggest that future work should focus on the effect of an automated generation of an optimal underlying scan dataset for a given task compared with manual curation of high-quality contours and varied datasets.

This work has several limitations. It is limited to the liver and may not be generalizable to smaller structures that have greater shape variability and less well-defined borders, such as tumors. Thus, we expect that, at a minimum, increasing the dataset size requirements we tested here will be needed to capture the variability of those structures. However, we expect that our results on liver images can be generalized to other whole-organ structures (lung, heart, kidney, etc.) because such structures have well-defined borders. Future studies should include datasets with multiple organ sites to validate further the generalizability of their results across fully withheld datasets. Although the large size of our liver dataset is a strength of this work, our dataset is still an order of magnitude smaller than many natural imaging datasets. Thus, future studies should bridge the gap in dataset sizes between medical and natural imaging. Future work should also consider other segmentation methods.

Although nnU-Net models offer several advantages, including state-of-the-art out-of-the-box performance and an automated configuration pipeline, those advantages may preclude generalization of these results to other architectures (Hatamizadeh et al., 2021; Roy et al., 2023) and less structured pre- or postprocessing pipelines.

Our results support that medical image datasets and segmentation do not follow the same rules as natural imaging regarding the prioritization of data quantity over quality. These findings were illustrated by smaller, highly curated models performing better in terms of DSC and surface DSC with 2mm margins than models trained on much larger, mixed-curation datasets. This observation has multiple potential explanations: the inherent differences in standardization and variability between medical and natural imaging, the different effects of 2D and 3D images, and, most notably, the much higher variability of contour quality in medical imaging than natural imaging because of a barrier of knowledge within medical imaging and radiologists’ consideration of non-imaging information.

## 5. Conclusion

We have shown that both approaches, focusing on a highly curated, smaller, higher-quality dataset or a mixed-curation, larger dataset, have distinct advantages and use cases to prioritize quantity or quality in dataset generation. For targeted use cases, a smaller, highly curated dataset offers equivalent performance to much larger, mixed-curation datasets. For generalizability larger mixed-curation datasets offer significantly better performance over smaller, highly curated datasets. In future work, we plan to validate our findings in other organ sites, tumors, and other imaging modalities.

## Data Availability

The BCTV data is available at https://www.synapse.org/Synapse:syn3193805/wiki/89480. The institutional datasets presented in this study are available upon request, in accordance with IRB protocol. Requests to access the datasets should be directed to K.K.B.

## Acknowledgments

This study was funded by the US National Institutes of Health and the US National Cancer Institute (grants 1R01CA235564, P30CA016672, and P01CA261669). Research reported in this publication was supported in part by resources of the Image Guided Cancer Therapy Research Program at The University of Texas MD Anderson Cancer Center, the Helen Black Image Guided Therapy Research Fund, the Image Guided Cancer Therapy Research Program at The University of Texas MD Anderson Cancer Center through a generous gift from the Apache Corporation, the Tumor Measurement Initiative through the MD Anderson Strategic Initiative Development Program (STRIDE). We gratefully acknowledge support from the Khalifa Foundation, the Andrew Sabin Family Fellowship, the Center for Radiation Oncology Research, the Sheikh Ahmed Center for Pancreatic Cancer Research, institutional funds from The University of Texas MD Anderson Cancer Center, GE Healthcare, Philips Healthcare, and Cancer Center Support (Core) Grant P30 CA016672 from the National Cancer Institute to MD Anderson. Dr. Eugene Koay was also supported by the NIH (U54CA210181-01, U01CA200468, and U01CA196403), the Pancreatic Cancer Action Network (14- 20-25-KOAY, 16-65-SING), Project Purple, and the Radiological Society of North America (RSD1429).

## Data availability and code availability

The BCTV data is available at https://www.synapse.org/-!Synapse:syn3193805/wiki/89480. The institutional datasets presented in this study are available upon request, in accordance with IRB protocol. Requests to access the datasets should be directed to K.K.B. Python code used for analysis and model training will be made available upon publication.

