## Supplemental Tables for "Quality versus quantity of training datasets for artificial intelligence–based whole liver segmentation"

### Slide 1
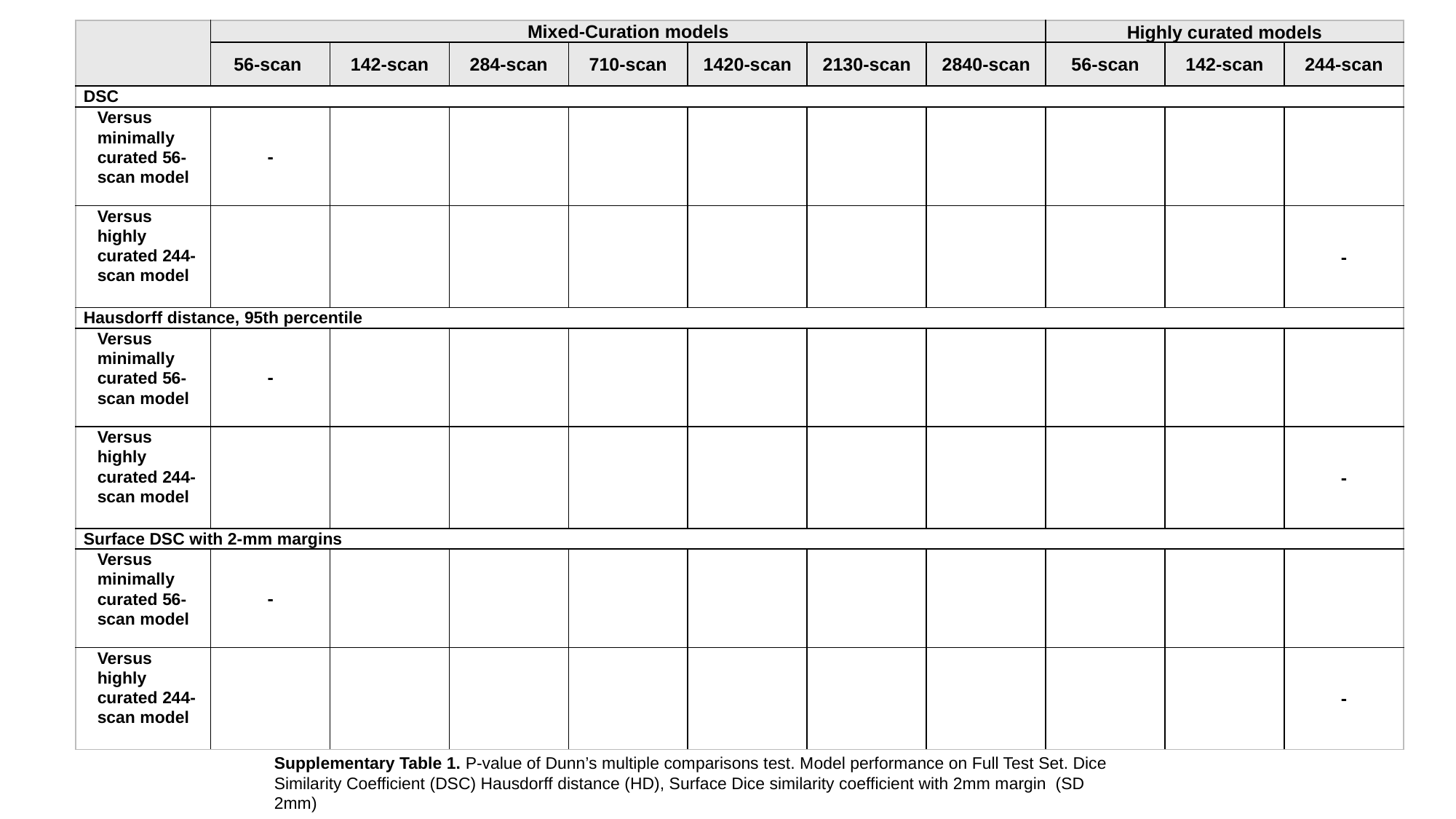

Supplementary Table 1. P-value of Dunn’s multiple comparisons test. Model performance on Full Test Set. Dice Similarity Coefficient (DSC) Hausdorff distance (HD), Surface Dice similarity coefficient with 2mm margin (SD 2mm)

### Slide 2
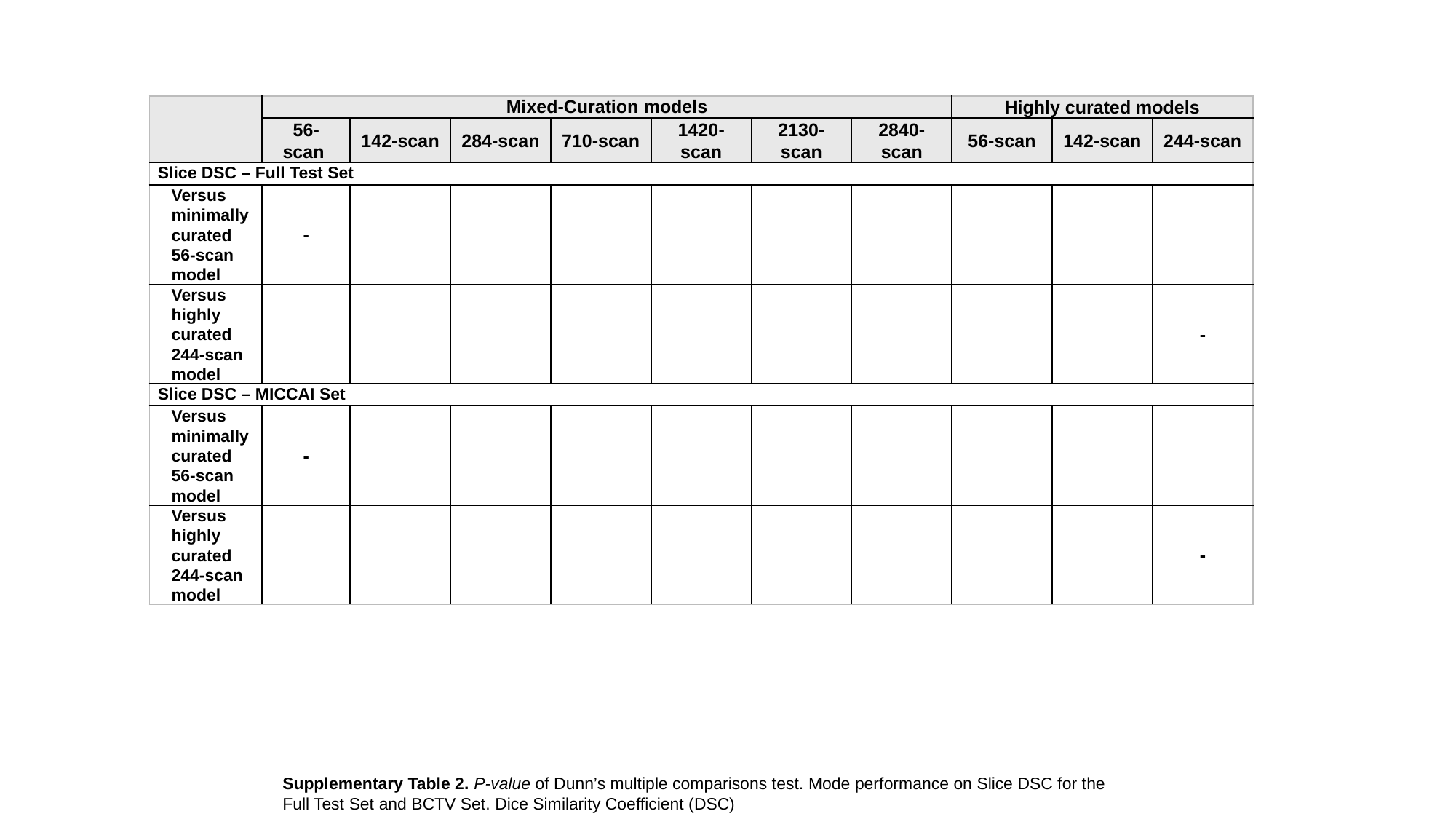

Supplementary Table 2. P-value of Dunn’s multiple comparisons test. Mode performance on Slice DSC for the Full Test Set and BCTV Set. Dice Similarity Coefficient (DSC)

### Slide 3
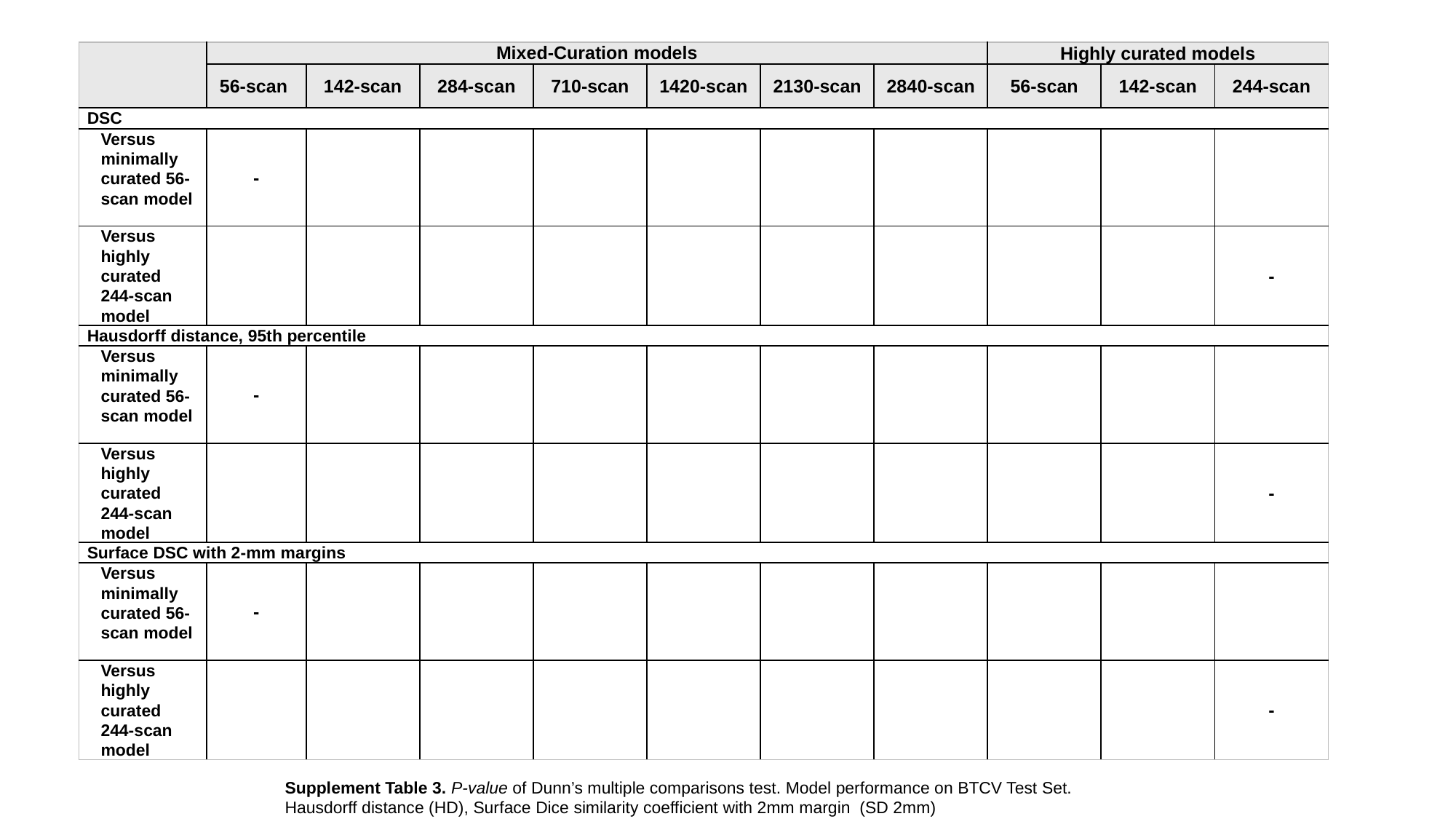

Supplement Table 3. P-value of Dunn’s multiple comparisons test. Model performance on BTCV Test Set. Hausdorff distance (HD), Surface Dice similarity coefficient with 2mm margin (SD 2mm)
